# Higher risk of COVID-19 hospitalization for unemployed: an analysis of 1,298,416 health insured individuals in Germany

**DOI:** 10.1101/2020.06.17.20133918

**Authors:** Nico Dragano, Christoph J. Rupprecht, Olga Dortmann, Maria Scheider, Morten Wahrendorf

## Abstract

**Background:** Previous research on infectious disease has revealed that infection risk as well as the severity of diseases is related to income and poverty. In this study we investigate if unemployed persons have a higher risk to become hospitalized with a COVID-19 diagnosis compared with employed persons.

**Methods:** We used routine data on hospitalizations in a study population of 1,298,416persons between the ages 18 and 65 who were enrolled in a German health insurance and who were active on the labour market (either employed or unemployed). Hospital diagnosis of COVID-19 (ICD-10-GM U07.1 and U07.2) were reported on a daily basis from 01.01.2020 to 04.06.2020. We studied if the rate of persons hospitalized with a COVID-19 diagnoses differed by employment situation. Logistic regression models comparing employed with short- and long-term unemployed were calculated adjusting for age and sex.

**Results:** In total, we observed 1,311 persons who were hospitalized, corresponding to a rate of 100.98 cases per 100.000 in our study population. Rates varied between the groups in different employment situations with lowest rates for employed and highest for long-term unemployed. Odds ratio for a hospitalization was 1.84 (1.64 - 2.07) for long-term and 1.18 (0.75 - 1.85) for short-term unemployed compared with employed persons.

**Conclusion:** The results are in line with earlier (mainly ecological) studies from the USA and Great Britain which found social inequalities in hospitalization risk. The fact that differences exist in Germany, a country with a universal health care system, indicates socioeconomic differences in the COVID-19 pandemic exists across countries.

## Introduction

Socioeconomic differences are found for most diseases (e.g. diabetes, respiratory diseases, coronary heart disease or depression) where people in more advantaged socioeconomic positions (SEP; usually measured on the basis of income, education and occupation) are healthier than their disadvantaged counterparts (1, 2). Studies also confirmed socioeconomic differences for various infectious diseases, including the H1N1 pandemic in 2009/2010 and seasonal influenza (3–10). In these studies, a disadvantaged socioeconomic position was both associated with risks of infection and severity of the disease (i.e. hospitalization, need of intensive care, and mortality).

These findings are in line with a currently growing number of studies from the US and England that report socioeconomic difference in the COVID-19 pandemic (for an overview see (11) or (12)). A recent study from New York (13), for example, compared the number of SARS-CoV-2 infections between neighborhoods and found that numbers were generally higher in more deprived neighborhoods than in rich areas (same for COVID-19-related hospitalization and mortality) (14). The same pattern became obvious across England in an analysis by the Office for National Statistics (ONS) that compared number of COVID-19 deaths between more than 32.000 areas and their level of deprivation (measured by the national index for multiple deprivation). In addition to these studies (which are based on ecological data with different levels of aggregation) a small number of recent studies also rely on individual data. Analyses of the UK biobank data, for example, show associations between low income and the probability of hospitalization (15), and between educational qualifications and SARS-CoV-2 infections (16). Furthermore, official individual data from the ONS revealed clear differences of COVID-19 mortality by occupational positions (17).

Whether theses inequalities exist for other countries (with other welfare and health care systems) is unknown, largely because national health monitoring of several countries does not consider any indicator of socioeconomic position, or because nationwide indicators of regional deprivation are not available on a level of small areas (18). It is the core objective of the present paper to help filling this gap of empirical evidence for Germany, a country where the pandemic was accompanied by a comparatively lower number of infections, as well as lower fatality rates up till now (19). We will focus on severe cases requiring hospitalization and their associations with unemployment. Unemployment is related to health in several ways, e.g. via psychosocial stress or poverty-related unhealthy behavior and is a known risk factor for acute and chronic diseases (20, 21). As underlying health conditions increase the likelihood of a severe COVID-19 disease that requires hospitalization, we assume that unemployment increases the likelihood of COVID-19 hospitalization. To study this association, we rely on individual level data from a German health insurance with information on 1.298.416 insured men and women. As an indicator of a disadvantaged socioeconomic position we use information on whether individuals receive any form of unemployment benefits (see Methods for details).

## Methods

### Population

Our study uses data from the German statutory health-insurance, namely the “AOK Rheinland/Hamburg”. This is one of the largest health insurances carrier in Germany for individuals living across Rhineland (western part of the state North Rhine-Westphalia) and Hamburg. For this study, we rely on data of 2,799,119 persons who were insured at any time during the observation period from 01.01.2020 to 04.06.2020 and applied the following restrictions: First, we restricted the analyses to men and women between the ages of 18 and 65 at the beginning of the observation period (to avoid an age-bias). Second, we restricted the analyses to individuals who were active on the labour force. This both includes people who are currently working and those who are unemployed but healthy enough to be available for work. Finally, we excluded students, retired persons (incl. those in disability retirement), and persons who were neither employed nor job-seekers (e.g. non-working partners of employed insured). As of 11 June 2020, these restrictions leave us with a group of 1,298,416 individuals (573,863 women, 724,553 men) who were active on the labour market at the beginning of the observation period (either employed or unemployed), and who were between the ages 18 and 65.

### Measures

#### COVID-19 hospitalization

People who are insured via the German statutory health insurance system have free access to hospital care in case of medical need. In case of hospitalization the responsible doctor records main and secondary diagnoses and this data are transferred to the insurance on a daily basis. Hospitalization due to COVID-19 is indicated by the international WHO ICD-10-GM code U07.1 (laboratory confirmed) and U07.2 (symptom based). In the infrequent case that more than one hospitalization was reported, we only consider the first hospitalization. During the observation period of the present study 418 patients were hospitalized with a U07.1 diagnosis and 893 with a U07.2 diagnosis. For the analyses, “caseness” was defined if either type of diagnosis was recorded (1,311 cases). We also conducted additional sensitivity analyses, where all findings were recalculated with confirmed cases (U07.1) only.

#### Employment situation

As part of its routine data collection, the health insurance also collects information on the legal employment situation of their members using a standardized coding system. This allows to distinguish between “regularly employed”, “short term unemployed”, “special benefits for low-income employees/job-seekers”, and “long-term unemployment”. A regular employment means that the insured person is working either as an employee or as a self-employed. Short-term unemployment means that the insured person is a job-seeker who receives benefits from the regular German unemployment insurance (about 60% of the previous income, called “ALG I”). For unemployed persons who do not find a new job within a defined period (see discussion) a special welfare system exists which provides a more restricted unemployment benefit (not calculated on the basis of previous income; called “ALG II”). A fourth category are low-salary workers or short-term unemployed with an income below a minimum threshold who additionally receive benefits from the “ALG II” system (called “Aufstocker” or “Ergänzer” in Germany). By definition, the reception of unemployment benefit is strongly related to a lower income, specifically in case of long-term unemployment and must be considered as a reliable and valid indicator of poverty risk and a disadvantage socioeconomic position for the active working population.

#### Additional Variables

The analyses also included age and sex, mainly as possible confounders in multivariable regression models.

### Data protection and ethics

Data are part of the routine data collection of hospitals and insurance carriers. The sample was completely pseudonymized preventing any de-pseudonymization by small subgroups for each analytical variable. The evaluation of the data was carried out exclusively by the AOK Rhineland/Hamburg that owns the data. Only descriptive/statistical results were made available, which do not allow any conclusions about the insured persons. The analyses plan was approved by the data protection office of the AOK Rhineland/Hamburg.

### Statistical methods

A simple analytical approach was applied. The cumulative incidence of hospitalization due to COVID-19 during the observations period was the binary outcome in a logistic regression model. Odds ratios and 95% confidence intervals were calculated comparing – for the main analyses – persons in employment as a reference population with the other employment situation groups. The outcome has a low incidence and fulfills the rare disease assumption. The odds ratios should therefore approximate relative risks closely. All calculations were performed using SAS 7.15.

## Results

The study population of 1,298,416 insured persons included slightly more men than women (55.8% men, see Table 1). The large majority was employed (72.2%), and most unemployed were long-term unemployed. Short-term unemployment and special benefits were less common. In total there were 1,311 cases of hospitalization with a COVID-19 diagnosis. This corresponds to a crude rate of 100.97 cases per 100,000 in the study population. The rate of hospitalization was generally higher among men and among unemployed, with highest rates of hospitalization for long-term unemployed.

**Table 1:** Sample description, main study characteristics including number and rate of cases of COVID-19 hospitalization (ICD-10-GM U07.1 + U07.2)

|  | observations and<br>column-% OR mean age<br>and standard deviation | cases with COVID-19<br>hospitalization | COVID-19<br>hospitalization, rate per<br>100.000 population# |
| --- | --- | --- | --- |
| *Total* | 1,298,416 | 1,311 | 100.97 |
| *Gender* |  |  |  |
| Female | 573,863 (44.20%) | 516 | 89.91 |
| Male | 724,553 (55.80%) | 795 | 109.72 |
| *Age* | 42.05 (SD 12.74) |  | - |
| *Employment situation* |  |  |  |
| Employed | 936,828 (72.15%) | 798 | 85.18 |
| Short-term unemployed | 16,702 (1.29%) | 19 | 113.76 |
| Special benefit recipients | 36,763 (2.83%) | 38 | 103.36 |
| Long-term unemployed | 308,123 (23.73%) | 456 | 147.99 |

#unstandardized

The results were confirmed in multivariable regression models that also included age (linear and squared) and sex (see Table 2). In details, the adjusted model shows that the odds for being hospitalized was 1.84 times higher for long-term unemployed compared to employed persons (references group). Compared with employed persons odds were also higher for short-term unemployed (1.18 times higher) and special benefit recipients (1.31 times higher).

**Table 2:**
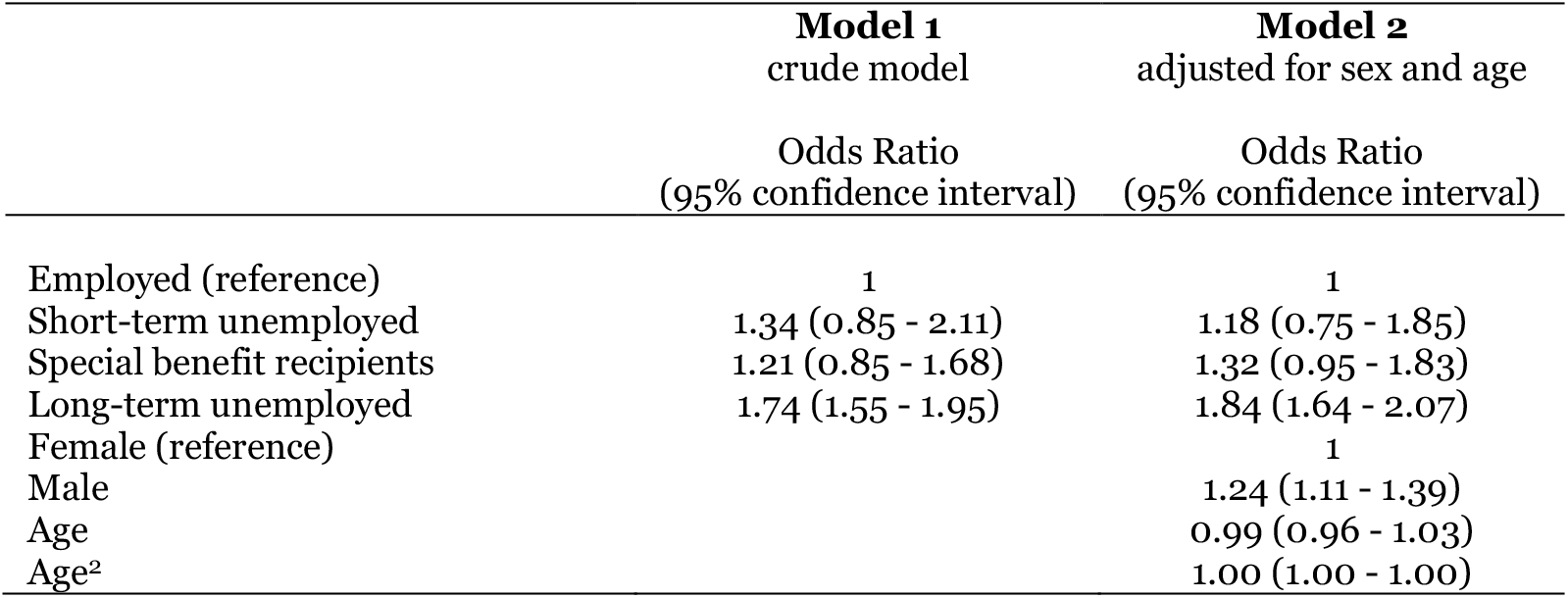
Results from multivariate logistic regression models on association between employment situation and COVID-19 hospitalization among 1298.416 health insured individuals

A separate analysis for men and women shows that findings are consistent for both genders, specifically in case of long-term unemployment (see Table 3). Slight differences were observed for short-term unemployment (stronger association for women) and for special benefit recipients (stronger associations) in men. These latter findings, though, deserve more detailed analyses (e.g. inclusion of interaction term) to draw conclusion about gender differences.

**Table 3:** Subgroup analyses by sex (logistic regression for risk of COVID-19 hospitalization, adjusted for age (linear and squared))

|  | **Female** | **Male** |
| --- | --- | --- |
|  | Odds Ratio<br>(95% confidence interval) | Odds Ratio<br>(95% confidence interval) |
| Employed (reference) | 1 | 1 |
| Short-term unemployed | 1.39 (0.66 – 2.95) | 1.07 (0.60 - 1.90) |
| Special benefit recipients | 1.20 (0.72 - 1.98) | 1.46 (0.95 - 1.06) |
| Long-term unemployed | 1.76 (1.47 - 2.12) | 1.88 (1.62 - 2.19) |

Results were generally confirmed in sensitivity analyses that were restricted to laboratory confirmed COVID-19 cases (ICD code U07.1), with the only exception of a less strong association in case of long-term unemployment (see Table 4).

**Table 4:**
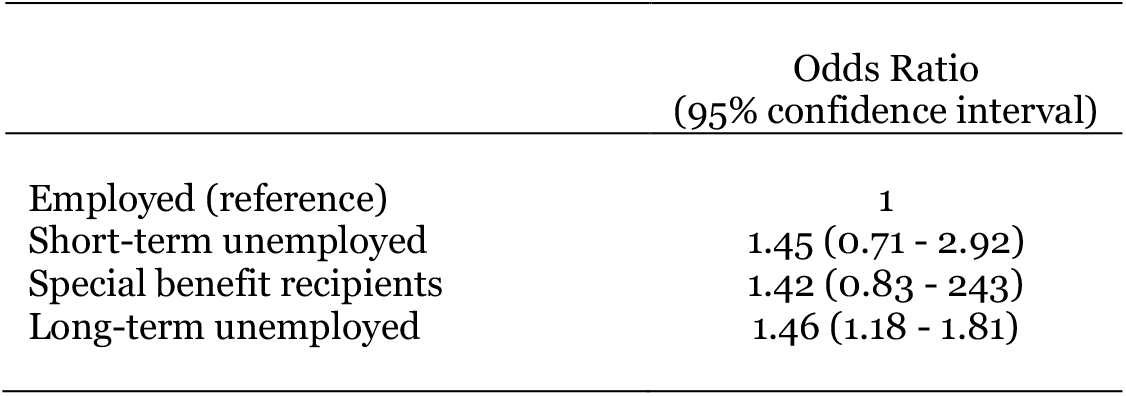
Sensitivity analyses using only the 418 laboratory confirmed cases (ICD-10-GM U07.1, n=418) as an outcome (results of logistic regression model adjusted for age and sex, total number of observations 1.297.523)

|  | Odds Ratio<br>(95% confidence interval) |
| --- | --- |
| Employed (reference) | 1 |
| Short-term unemployed | 1.45 (0.71 - 2.92) |
| Special benefit recipients | 1.42 (0.83 - 2.43) |
| Long-term unemployed | 1.46 (1.18 - 1.81) |

## Discussion

In this study of around 1,3 million German health insured we found a gradient in the risk of hospitalization with a COVID-19 diagnoses in short- and long-term unemployed compared with gainfully employed persons. This finding is in line with findings from previous epidemic outbreaks such as the H1N1 pandemic in 2008 and with recent reports from the COVID-19 pandemic (11, 12, 14, 17, 22–24). Reliable data on COVID-19 is still limited to a small number of countries and this first report from Germany – a country with universal welfare and health care access – adds new evidence for the existence of a social gradient in infectious disease including COVID-19.

While the evidence of health inequalities in the COVID-19 Pandemic is growing, one important challenge is to understand the reasons for these inequalities. Based on a framework by Quinn and colleagues, three potential explanations can be distinguished (22, 25): First, different living and working conditions may lead to *inequalities in exposure* to the virus (22). For example, ongoing studies confirm that people in more advantaged socioeconomic positions work less frequently in occupations with a high exposure risk. Similarly, people with lower income are more likely to the live in adverse environments (including crowded housing conditions or possible exposure in public transport). A second explanation refers to *inequalities in vulnerability*. Underlying health conditions are more frequent among socioeconomically disadvantaged groups, thus, leading to increased risk of infections and severe health outcomes (26, 27). Likewise, recent studies highlight the role of environmental pollution (e.g. air pollution) and their impact on underlying health conditions (28). It seems likely that disadvantaged population groups rather live in areas where pollutions levels are high (29). A third explanation are *Inequalities in care*. These include comparatively limited access to medical care (e.g. difficult access to medical facilities (30) or - as shown in a US-American study - less frequent opportunities for testing in disadvantaged areas (31)), but also differences in utilization (e.g. delayed symptom awareness and later help seeking behavior (32)). Exploring these potential pathways (and their relative importance for different outcomes and indicators of SEP) will surely be part of future studies.

For the present study, we assume that a higher vulnerability due to underlying health conditions is a plausible link. The prevalence of chronic conditions such as cardiovascular disease and of behavioural risk factors like smoking or obesity is higher for unemployed persons in many countries (21, 33) including Germany (34). Those conditions are important risk factors for a severe progression of a SARS-CoV-2 infection and higher rates among the unemployed may explain our findings (35). Regarding a differential exposure no data exists comparing workers with unemployed populations. Possibly, though, the risk of infection may be even lower for unemployed because of their lower mobility (i.e. commuting) and the fact that they are not exposed to the virus through social contacts with colleagues, customers or patients during work. It was not possible to account for differences in infection risk per se in this analyses because data was not yet available and future investigations combining risk of infection and risk of severe diseases are needed. The third explanation (inequalities in care) may be less important in our case. The German health care systems offers universal access to both ambulatory and hospital care. Utilization of health care, however, could have played a role as data from Germany shows that poor people avoid the utilization of health care – unless universal access – more often than more affluent persons (30, 36).

### Strengths and Limitations

We were able to study the complete population of insured people of the insurance carrier who meet the inclusion criteria which guarantees a high internal validity of the sample. The large number of observations and the reporting of diagnosis on a daily basis must be considered as further a strength of the analyses. Our measure of unemployment is also based on official standardized records which meant that the reliability of the main exposure variable was high. Yet, the present study has important limitations and calls for future studies. First, we used data from only one statutory insurance carrier for people living in specific regions of Germany, while there are several other carriers in Germany. Our study population may therefore be selective. Specifically, it is well-known that the health insurance we have used in this study does historically over-represent persons with unemployment. This does surely prevent to draw conclusions about the composition of the labour force in Germany (e.g. percentage of unemployed). But it is unlikely that it affects the reported associations between unemployment and hospitalization. Second, our study relies on data and an observation period that covers an early period of the COVID-19 pandemic with rather low number of infections from January to March 2020, and we may question if findings will be even stronger once the Pandemic has progressed. Next, administrative data includes only limited information on socio-demographic and socio-economic position.

An important limitation is the lack of information on diagnosis of COVID-19 treated in outpatient care. It is well documented that the majority of persons with a COVID-19 diagnosis is not hospitalized and it must be expected that a large number of COVID-19 diagnosis was unmeasured in our study and no conclusions about overall infection rate can be made. Missing information on the timing of the diagnosis is also a problem. Detection bias may have occurred as unemployed are more often hospitalized due to other chronic conditions and that diagnosis were more likely to be made in the hospital as part of routine care. Thus, future analyses also need to consider different outcomes, such as infection risk, use of intensive care or mortality among those who are infected.

Another limitation is that we used a relatively simple modelling approach (logistic regression with cumulative incidence as an outcome) which does not allow to account for the (small) number of persons who died for other reasons than COVID-19 during the observation period or who changed their employment situation.

Finally, it was not possible to adjust for the duration of unemployment as we defined unemployment using official coding of insurance status. In Germany, short-term unemployed receive benefits from the unemployment insurance, while long-term unemployed are enrolled in another system of benefit. The time after which a person switches from short- to long-term is legally defined and dependent of age and duration of the payment of contributions to the unemployment insurance. It lasts between 6-24 Months and there is therefore a certain heterogeneity within the groups.

In conclusion, this study delivers first empirical evidence of differences in COVID-19 hospitalization by employment situation for Germany. If further supported by future studies with alternative indicators of socioeconomic position together with different outcomes of morbidity (incl. risk of infection, risk of intensive care) and mortality, this underlines the importance of socioeconomic factors. These factors should be considered – together with age and underlying health conditions – as important factors to identify high-risk groups and to develop infection control measures (18). In addition, our study highlights the importance to improve the availability of data in Germany and to collect information on socioeconomic factors to allow for studying socioeconomic difference during the COVID-19 pandemic.

## Data Availability

Administrative data used are protected by law and analyses must be performed by the data owner AOK Rhineland/Hamburg

